# Estimating Spatial and Temporal Patterns of Residential Power Outages in Massachusetts from 2013 to 2022 Using Public Records

**DOI:** 10.1101/2025.07.08.25331086

**Authors:** Chad W. Milando, Muskaan Khemani, Allison M. James, Jianna Correi-Silva, Jill Collins, Madeleine K. Scammell, Mary D. Willis, Jonathan I. Levy, Amruta Nori-Sarma

**Affiliations:** Boston University School of Public Health, Department of Environmental Health; Boston University School of Public Health, Center for Climate and Health; Boston University, Center for Health Data Science; Conservation Law Foundation and Tufts University, School of Engineering; Boston University School of Public Health, Department of Epidemiology; Harvard T.H. Chan School of Public Health, Department of Environmental Health; Harvard T.H. Chan School of Public Health, Center for Climate, Health, and the Global Environment (C-CHANGE)

**Author notes:** corresponding author, 715 Albany Street Boston MA, 02118.

**Keywords:** power outage, exposure, public utilities, energy, electricity

## Abstract

Power outages are a growing threat to human health. Extreme weather events and strains on the electrical grid can cut off access to critical health-supporting medical equipment and create immense stress. Outages have been associated with premature mortality, pregnancy complications, mental health emergency room visits, and other health outcomes. However, the full scope of health impacts related to power outages have not been quantified, as outage data with high spatial and temporal resolution are not widely available. In this work, we present a methodological framework for constructing a longitudinal, high spatial and temporal resolution dataset of power outages using public records. We apply this method to the Commonwealth of Massachusetts, extracting and synthesizing data from daily town-level reports submitted by electricity providers to the Massachusetts Department of Public Utilities from 2013 to 2022. For each town-day, we calculated the fraction of electrical circuits experiencing a power outage and classified this into a tertile categorical variable: Mild, Moderate, or Severe. Across the state, towns experienced an average of 0.05 to 1.7 outages of each category per month, with peaks in March, July, and October. Mild outages were most related to equipment failure or planned maintenance, whereas Moderate and Severe outages were most related to tree interference (35.6% and 50.9% of outages, respectively). Developing suburbs and rural towns experienced the highest frequency of Severe outages, while inner core communities, regional urban centers, and towns with a high density of environmental justice populations experienced more frequent Mild outages. Our dataset reinforces substantial spatial and temporal heterogeneity in power outages, and our methods provide a framework for building similar datasets for future analyses of population vulnerability and health impacts associated with power outages in other states where curated datasets are not available.

## Introduction

Urbanization and population growth across the US over the past several decades have led to increasing strains on critical infrastructure, including power supply. As climate change increases the frequency, duration, and severity of extreme weather events, power outages caused by disruptions to the electrical grid are expected to increase as well.^1^ Aging electrical grid infrastructure also contributes to increased frequency of power outages; today, over 70% of the US electrical grid is over 25 years old,^2^ increasing the vulnerability of the system to extreme weather events among other outage causes. Power outages are thus a large and growing threat to human health, as both medically vulnerable persons and healthcare systems rely on electricity for medical care provision. Lack of electricity can cut off access to medical equipment and create immense stress,^3,4^ which can exacerbate existing medical conditions or even lead to health impacts directly.^1,5^

The majority of 8+ hour power outages co-occur with extreme weather events,^6,7^ and this type of outage were more than twice as frequent from 2014 to 2023 than from 2000 to 2009.^8^ Extreme weather can also add strain on the aging electrical grid infrastructure, especially as electrification efforts and greater use of air conditioning during increasingly hot summers increase electricity demand, leading to adverse health impacts.^9–11^ For example, when Hurricane Irma hit Florida in September 2017, loss of electricity was associated with increased adjusted odds of mortality over the following 7 to 30 days.^12^ Several counties in New York City experienced a rise in mental health emergency room visits after power outages associated with Hurricane Sandy in October 2012.^13^ Power outages occurring after Hurricanes Sandy (in New York State, 2012) and Maria (in Puerto Rico, 2017) were associated with an increased risk of adverse pregnancy outcomes.^14,15^ The impacts of power outages may potentially increase as extreme weather events increase in frequency, severity, and duration.^1^ As power outages become more frequent, prolonged, and severe, high-resolution data are essential for effective emergency planning and protecting population health.

Fully characterizing the health impacts of power outages, and populations disproportionately burdened by those impacts, requires power outage exposure data with high spatial and temporal resolution over a long duration. These data are not widely available across the US, limiting large-scale nationwide studies that can identify environmental justice communities with respect to outage exposures as well as studies of the compounding health effects of outages that co-occur with extreme weather events. A variety of exposure datasets have been used to date to explore exposure inequities and health impacts, but each has limitations (**Table 1**). Many studies utilize data from single power outage events, including in New York,^13,14,16^ Florida,^12^ Texas,^17^ and Puerto Rico.^15^ These single outages provide useful insights about short-term health impacts, but repeated exposure to power outages may also have additive or multiplicative health impacts not captured by studies of single events. Although national-scale data exist at the county-level,^6,7,17,18^ higher spatial resolution data (i.e., below the county-level) are needed to identify the specific populations that are likely to be most affected,^19,20^ especially for rural communities where counties may cover a large area with heterogeneity in the vulnerability of local residents.

**Table 1.** Overview of power outage exposure metrics used in studies in the United States.

| Reference | Metric | Spatial Span | Spatial Resolution | Temporal Span | Temporal resolution |
| --- | --- | --- | --- | --- | --- |
| This manuscript | Tertiles of % of circuits affected | Massachusetts | Town-level | 2013-2022 | Daily |
| Anderson & Bell, 2012 | Day has outage (yes/no) | New York, NY | City | Aug. 14–15, 2003 | Daily |
| Lin et. al., 2016 | Number of house total blackouts | Southern New York State | County | Oct. 28, 2012, to Nov. 9, 2012 | Daily |
| Dominianni et. al., 2018 | Day has outage (yes/no) | New York, NY | Electric-grid networks <sup>a</sup> | 1997-2014 | Daily |
| Zhang et. al., 2020 | % customers affected, and Binary variable: yes/no % customers affected > 50<sup>th</sup> percentile | New York State | Operating division <sup>b</sup> | 2001-2013 | Daily |
| Lin et. al., 2021 | Binary variable: yes/no % customers affected > 50<sup>th</sup> percentile | New York State | Operating division <sup>b</sup> | 2001-2013 | Daily |
| Sheridan et. al., 2021 | Binary variable: yes/no % customers affected > 50<sup>th</sup> percentile | New York State | Operating division <sup>b</sup> | 2005-2013 | Daily |
| Skarha et. al., 2021 | Day has outage (yes/no) | Florida | Nursing homes | Sept. 2017 | Daily |
| Xiao et. al., 2021 | Binary variable: yes/no % customers affected > 50<sup>th</sup> percentile | New York State | Operating division <sup>b</sup> | Oct. 29, 2012, to Nov. 27, 2012 | Daily |
| Deng et. al., 2022 | Binary variable: yes/no % customers affected > 75<sup>th</sup> percentile | New York State | Operating division <sup>b</sup> | 2002-2018 | Daily |
| Do et. al., 2023 | % customers affected | Nation-wide | County | 2018-2020 | Hourly |
| Flores et. al., 2023 | % customers affected | Texas | County | February 2021 | Hourly |
| Flores et. al., 2024 | % customers affected | New York State | Operating division <sup>b</sup> | 2017-2020 | Hourly |
| Pradhan et. al., 2024 | Avg # of households experiencing an outage > 12 hours | Minnesota (one electricity provider) | Block group | 2017-2022 | 2-year averages |
| Northup et. al., 2024 | % customers affected, cumulative over 36 hours | New York State | Operating division <sup>b</sup> | 2017-2020 | Daily |
| Do et. al., 2025 | % customers affected by outages 8+ hours in length | Nation-wide | County | 2018-2020 | Hourly |

<sup>a</sup> An electric grid network roughly overlaps with collections of ZIP-codes. <sup>b</sup> An operating division is approximately equal to a ZIP-code in size.

While nationwide studies are limited, prior work has demonstrated successful implementation of high spatial and temporal resolution datasets of power outages, for example in the State of New York. New York data with spatial resolutions of grid-network (i.e., groups of ZIP codes) and operating division (i.e., individual ZIP codes) the have been used to quantify multiple health impacts related to power outages: all-cause mortality during cold-related power outages;^21^ injury emergency department visits during power outages co-occurring with high wind events,^22^ ice and snow-storms,^23^ and floods;^22^ pregnancy complications for power outages that follow a hurricane;^14^ emergency department visits specifically for chronic obstructive pulmonary disease;^24^ and pediatric injury hospitalizations.^25^ These data were also used to examine whether power outages were associated with elevated levels of cardiovascular hospitalizations among Medicare fee-for-service recipients, with null associations reported.^26^ In the absence of a curated state-level dataset, public records may be leveraged to characterize the spatial and temporal variation in power outages.

For the state of Massachusetts, community feedback has indicated that there is a high priority need for power outage data to inform health protective strategies among vulnerable residents. We address the gap in available data for power outage exposures in MA by providing a methodology for creating a power outage exposure dataset from public records and demonstrate its capacity to characterize exposure to power outages at high spatial and temporal resolution across 10 years. Here, we created a daily power outage database at the town-level for the Commonwealth of Massachusetts using data from the years 2013-2022. We then analyzed these data and converted the fraction of a town’s electrical circuits that are experiencing a power outage into a categorical variable with three levels: Mild, Moderate, and Severe, to facilitate continued research on disproportionately impacted communities and potential health consequences of power outages.

## Methods

### Power outage exposure data

We gathered power outage data from public records of the Massachusetts Department of Public Utilities. Since 2001, utility companies in Massachusetts are required to report significant interruptions in services through quarterly reports. Significant outages are defined throughout as “one that: (1) affects 500 or more customers and is likely to extend for one hour or more; and/or (2) affects a critical facility; and/or (3) has an expected duration of 500 customer hours even if it affects fewer than 500 customers.”^27^ For these analyses, we excluded outages that affect critical facilities (i.e., buildings that would cause a disruption in critical public safety if affected by an outage, such as hospitals, police and fire stations, airports, and others), as their patterns of outages are unique and they often have emergency generators on site.^28^

Outage records at the individual circuit level are listed in two separate documents for each fiscal year with different reporting frequencies: Annual Service Quality Reports listed existing circuits for each utility company each year, and quarterly Significant Outage Reports detailed when specific circuits were out of operation. These reports were primarily in Excel or PDF format. For data provided in PDF format, a Python script was utilized to obtain data from tables using the “pdfplumber” package^29^ and automatically export it to a CSV file. Different scripts were developed for each electricity distributor to accommodate for different table formats, missing or duplicated data for certain towns or months, spelling errors, and irregular spacing. For each year, the Significant Outage Reports documents were catalogued to identify the documents that contained information on outage day, duration, town attributed to each circuit, circuit number, number of customers affected, and cause. Similarly, Service Quality Reports documents were catalogued to identify documents containing number of customers served, town, and circuit number. Information included in reports for each outage consisted of the feeder’s name, outage cause, event total customer hours, event total customers affected, and event duration, with varying degrees of missingness. Outage and customer information data were then joined together at the circuit level to contextualize outages and be able to estimate the percentage of customers exposed. Estimating the number of customers served in each town required the assumption that a circuit serving multiple towns served an equal number of customers in each town. For outages that spanned multiple days, an even distribution of outage time between days was assumed. Written causes for outages (i.e., free text fields) were manually sorted into categories to enable trend analyses (**Table S1**). Of 351 cities and towns in Massachusetts, 310 reported outages between 2013 and 2022 (**Table S2**). Municipalities without reported information either contained municipal light plants that did not submit outage data to the state (n = 49 by 2022) or had no reported outages.

### Town classification data

We used two metrics to classify towns based on the potential underlying infrastructure and demographic vulnerability. The Metropolitan Area Planning Council, a government research organization that enables regional collaboration in Massachusetts among lawmakers and municipal stakeholders, created a statewide community types dataset which labels towns as Inner Core, Regional Urban Centers, Maturing Suburbs, Developing Suburbs, and Rural Towns (**Table 2**). We also used the Massachusetts definition of an Environmental Justice community as one where greater than 50% of the census block groups in a town meet one or more of a set of classifications (**Table 3**).

**Table 2.** Metropolitan Area Planning Council community types.

| Community type | Description |
| --- | --- |
| Inner Core | • High-density, built-out neighborhoods near the urban core, featuring a mix of apartments, multifamily homes, and some single-family housing.<br>• New development is limited and primarily occurs through infill, redevelopment, or expansion of existing structures, including conversions from industrial use.<br>• Communities are shaped by historical urban development patterns, including recovery from mid-20th century disinvestment and suburban flight.<br>• Populations are often diverse—especially in core neighborhoods—with many minority and immigrant residents; population trends range from stable to slightly declining, partly due to shrinking household sizes. |
| Regional Urban Centers | • Typically have over 70,000 residents, with diverse housing types—mostly multifamily—centered around a dense downtown core and surrounded by suburban-style neighborhoods.<br>• Often largely built out, with limited vacant developable land; however, some may still have greenfield areas at the periphery.<br>• New development primarily occurs through redevelopment, infill, and industrial-to-residential conversions, with some peripheral cities also experiencing suburban expansion.<br>• These communities tend to have a modest or declining tax base per capita and are either growing slowly, stable, or recovering from past population loss. |
| Maturing Suburbs | • Mid- to low-density suburban communities, primarily composed of owner-occupied single-family homes on ¼ to 1-acre lots.<br>• Nearly or approaching full buildout, with limited vacant developable land (typically less than 15–20% of land area).<br>• New residential growth occurs mainly through infill, teardowns, small-scale redevelopment, and occasional greenfield development.<br>• Populations are generally stable or growing at a modest pace. |
| Developing Suburbs | • Low- to moderate-density towns with substantial vacant developable land (over 25–35% of total area), offering significant room for growth.<br>• Growth is driven by conventional low-density subdivision development on previously undeveloped land.<br>• Maturing towns often feature a defined mixed-use center with compact neighborhoods, while country suburbs lack a central core and remain more dispersed in layout.<br>• Both types are experiencing rapid population and household growth, with increasing conversion of land to residential use |
| Rural Towns | • Very low density communities with no significant town center and scattered “farmstead” settlements; very few subdivisions; very limited economic development<br>• Very large amounts of vacant developable land (>40% of total town area is vacant & developable)<br>• New growth: small amounts of scattered residential development (average below 15 acres/year)<br>• Population less than 2,500 and growing slowly |

Note: for additional information, see https://www.mapc.org/wp-content/uploads/2017/09/Massachusetts-Community-Types-Summary-July_2008.pdf

**Table 3.** Definition of an Environmental Justice neighborhood in Massachusetts, a neighborhood where one or more of the following criteria are true.

| Criteria | Description |
| --- | --- |
| (a) | Annual median household income is 65 percent or less of the statewide annual median household income |
| (b) | Minority populations <sup>a</sup> make up 40 percent or more of the population |
| (c) | 25 percent or more of households identify as speaking English less than "very well" |
| (d) | Minority populations make up 25 percent or more of the population and the annual median household income of the municipality in which the neighborhood is located does not exceed 150 percent of the statewide annual median household income. |

<sup>a</sup> Minority is defined as 2020 Census Redistricting Data: Table P2: HISPANIC OR LATINO, AND NOT HISPANIC OR LATINO BY RACE. For more information see: https://www.mass.gov/info-details/environmental-justice-populations-in-massachusetts

### Data analysis

To assess exposure to power outages, we first identified the daily number of circuits affected in each town. From this, we calculated the daily percentage of circuits affected. Next, to enable characterization of outage frequency and outage severity, we created a categorical variable (defined by tertiles of the percentage of circuits affected) that represented different levels of outage severity. For each category, we also calculated the fraction of outages attributed to different causes. Finally, we merged the town classification data for each town with the outage data to explore the relationships between town classification, outage severity, and outage days. We analyzed temporal patterns in daily outages at the county level using a quasi-Poisson model including a random effect for county and controlling for year and month. This method permits examination of the spatial pattern of outages of varying severity and allows us to identify trends by important predictors of vulnerability.

## Results

We created a categorical exposure variable for outage severity using tertiles of the percentage of circuits affected on a particular day (**Figure 1**), noting that the number of circuits per town was correlated with town population (Pearson correlation coefficient = 0.92). The tertile categories were Mild outages (less than 3.3% of a town’s circuits were affected on a particular day), Moderate outages (3.3 to 8.5% of a town’s circuits affected), and Severe outages (greater than 8.5% of a town’s circuits affected).

**Figure 1.**
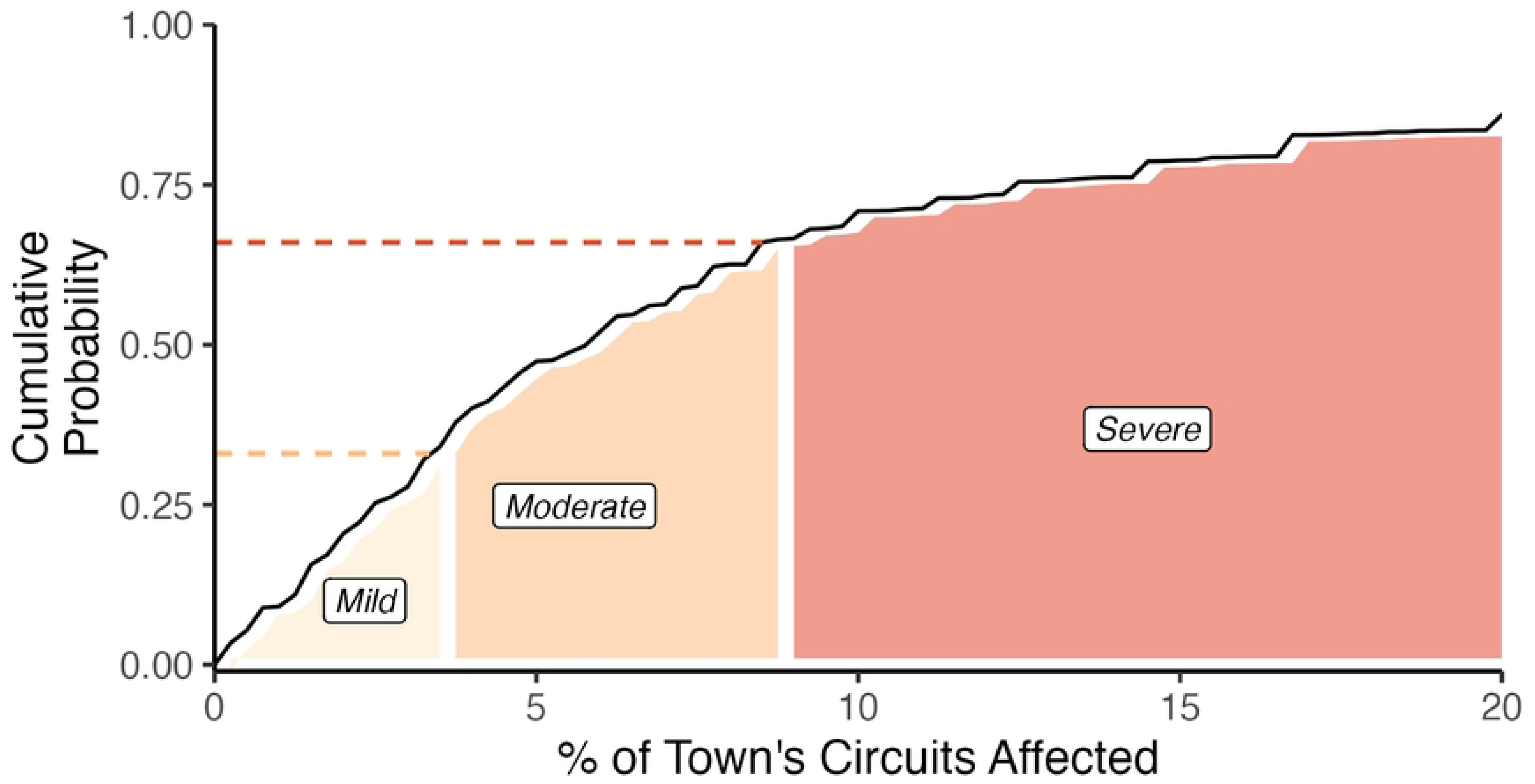
Outage severity categories - Mild, Moderate, and Severe - as a function of the percentage of a town’s circuits affected. The tertile break points were 3.3% (Mild to Moderate) and 8.5% (Moderate to Severe).

From 2013 to 2022, towns had an average of 0.05 to 1.70 outages of each severity category per month (**Figure 2**). Regression modeling showed that March, July and October were the months where outage counts by county diverged most from the reference month (January) controlling for year-by-year trends. There was no year-by-year trend when controlling for month (**Table S3**).

**Figure 2.**
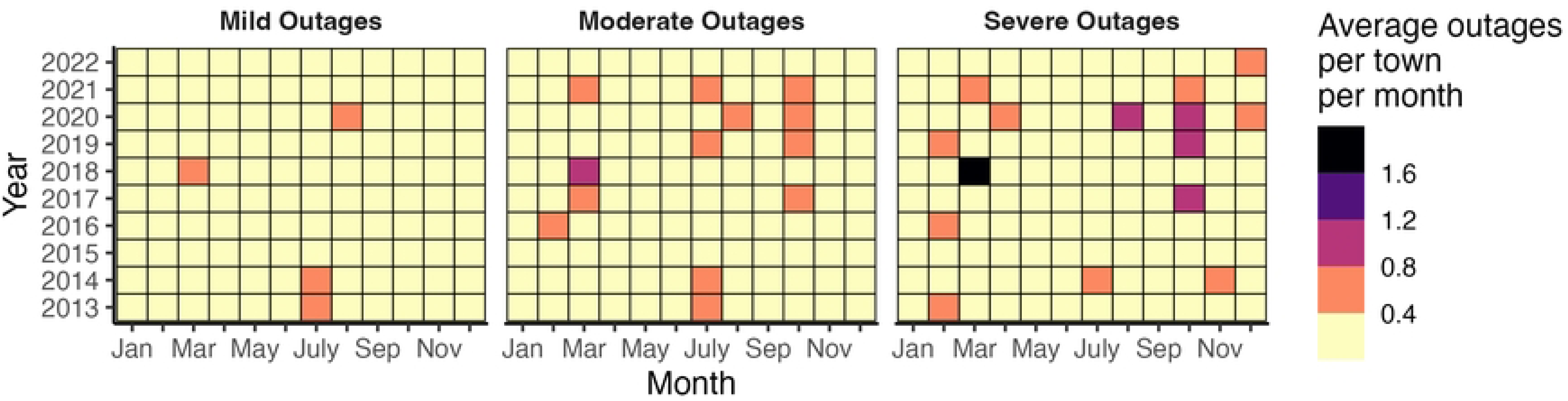
Average outages per town per month from 2013 to 2022, separated by Mild, Moderate, and Severe outages (defined by tertiles of the percentage of circuits affected on a particular day). The range of the scale is 0.05 to 1.70.

Outages of different severity had different labeled causes (**Figure 3**). For Mild outages, equipment failures were the most common label, followed by planned maintenance. For both Moderate outages and Severe outages, interference from downed trees was the most common labeled cause (35.6% and 50.9%, respectively).

**Figure 3.**
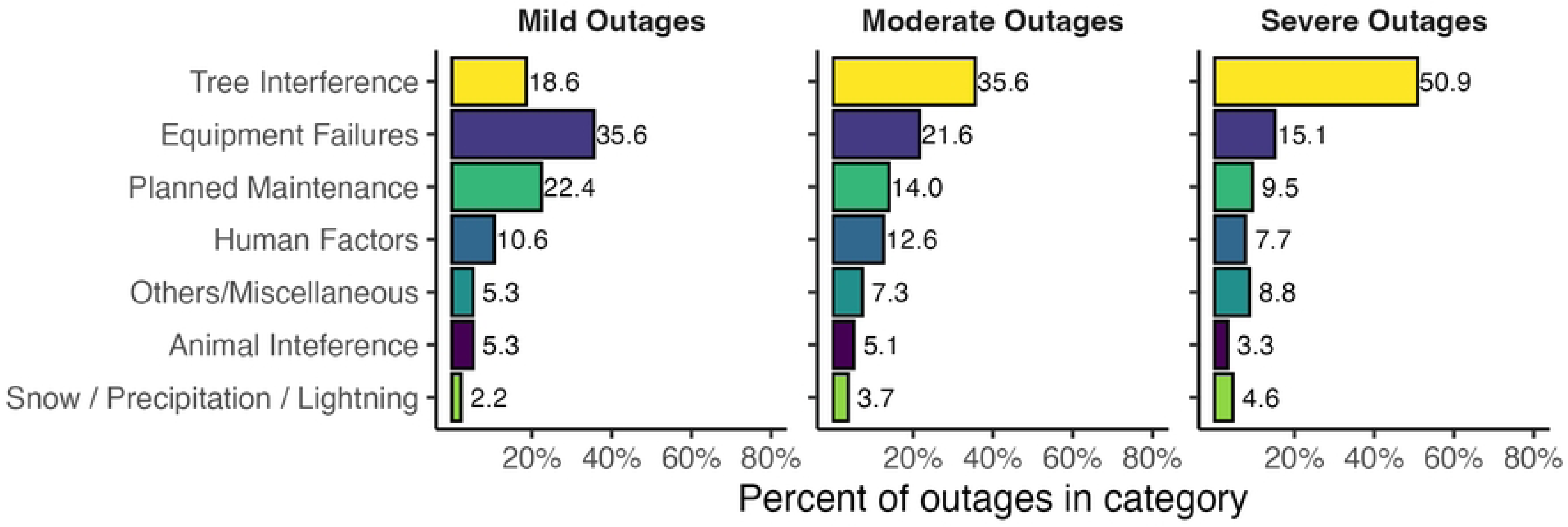
Percent of Outage-days from 2013 to 2022 attributable to different causes, stratified by severity of outages.

Labeled causes of outages also differed by season (**Figure 4**). For example, there was pronounced seasonality for Severe outages, with elevated outage rates in February, March, and October, driven by tree interference. In contrast, seasonality was more limited for Mild outages, with slightly higher outage rates in summer months given modest increases in equipment failures and animal interference. Across all severity types, tree interference had elevated contributions in late winter, summer, and mid-fall, but with large differences in relative contributions and the extent of seasonality.

**Figure 4.**
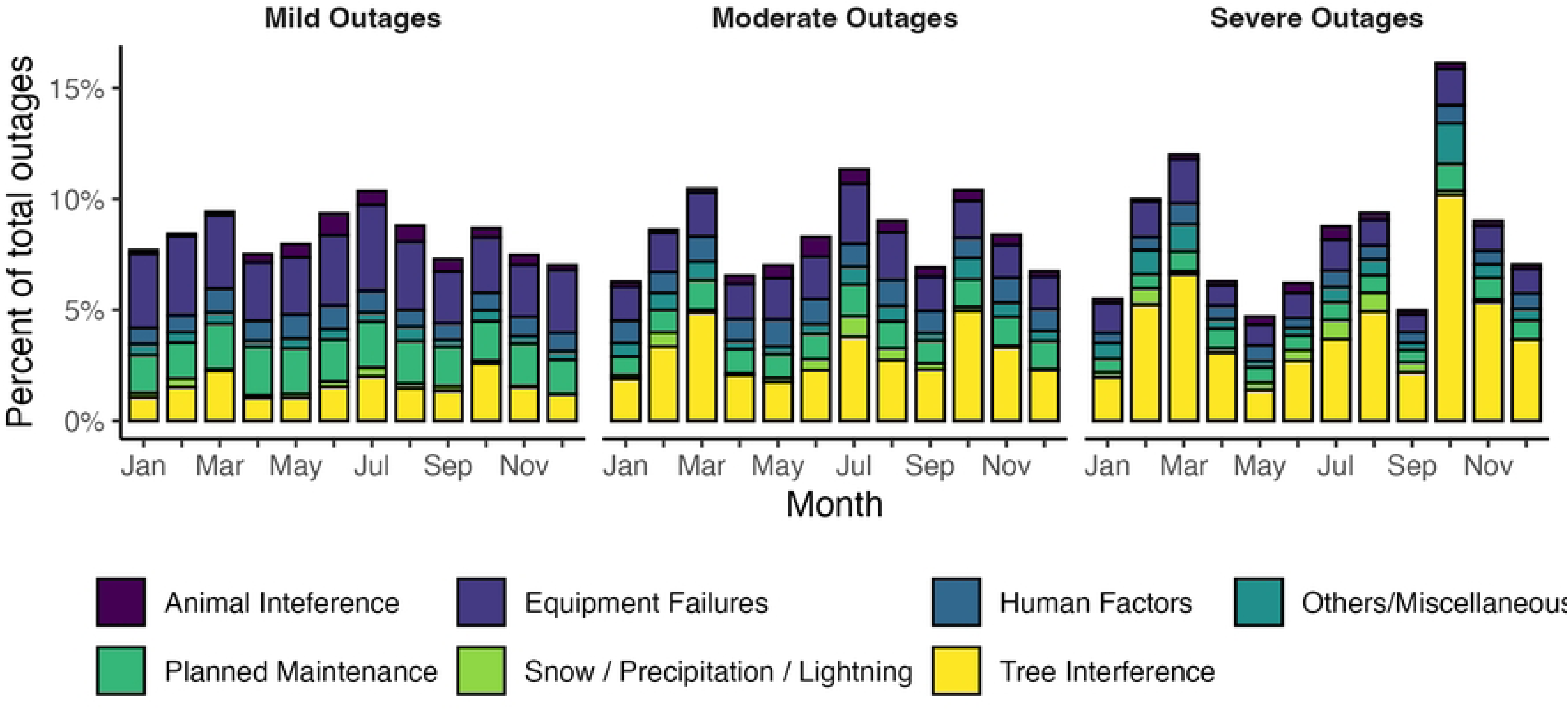
Monthly distribution of outages per town from 2013 to 2022, stratified by outage cause and severity (defined by tertiles of the percentage of circuits affected on a particular day).

We observed variation in outage severity and frequency spatially within counties, with the most heterogeneity in outage intensity occurring within counties in the western part of the state (**Figure 5**). These western counties contain cities and towns with fewer circuits, thus outages in just a few circuits will result in a large percentage of circuits affected.

**Figure 5.**
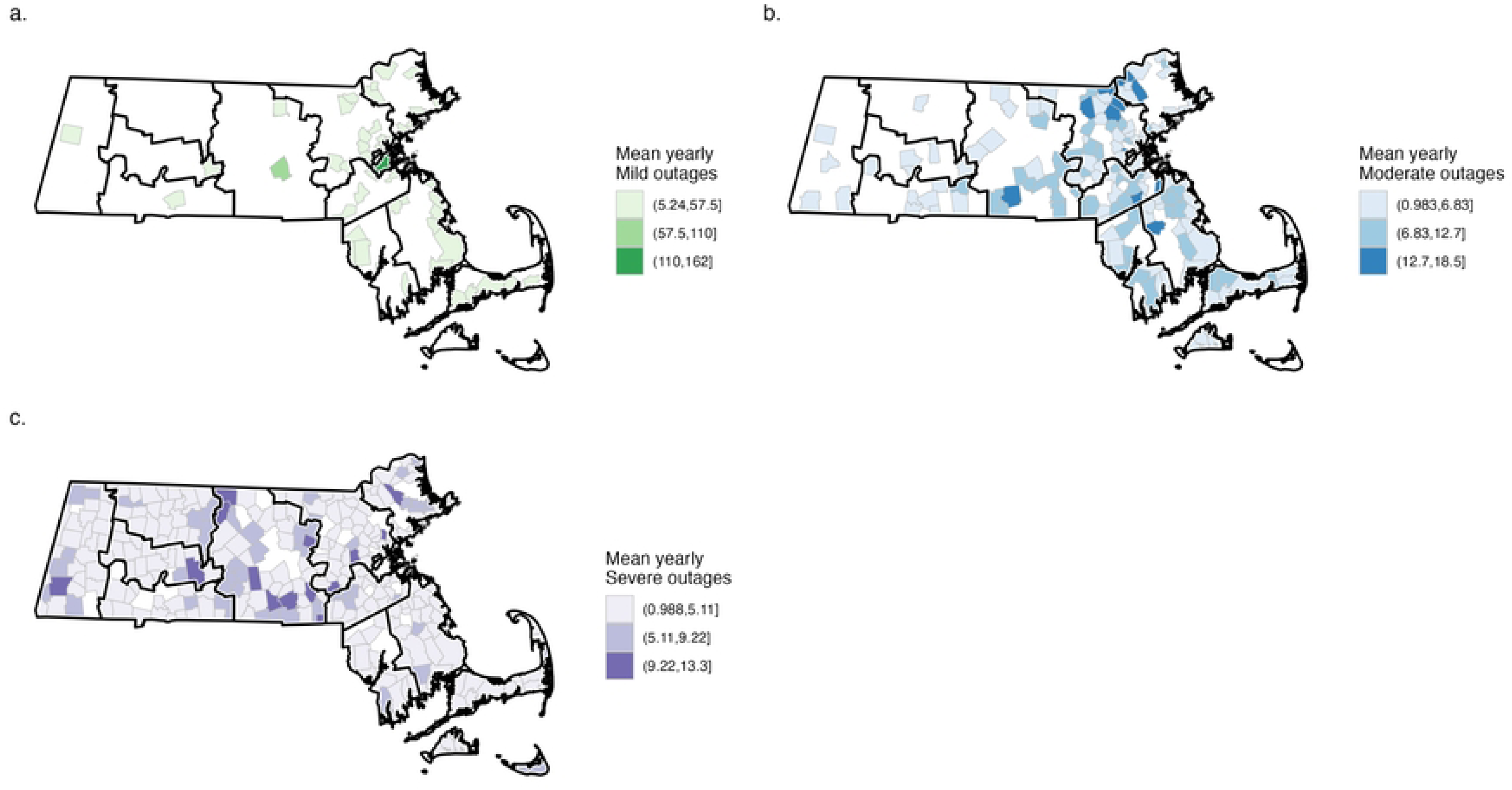
The spatial distribution of the yearly mean outages for (a) Mild, (b) Moderate, and (c) Severe outages. Towns are outlined in grey, and counties are outlined in black – towns not shown have no listed outages of that category.

We also observed systematic differences in outage frequency across community types (**Figure 6**). Mild outages most frequently occurred among towns classified as Regional Urban Centers and Inner Core (which contains the Boston Metro area), whereas Moderate and Severe outages were more common in Developing and Maturing Suburbs and Rural towns. Rural towns had no Mild outages – largely due to the low number of circuits in these towns. Similarly, in Massachusetts, Environmental Justice populations are most concentrated in Region Urban Centers and the Inner Core, and thus these cities and towns experienced a higher frequency of Mild outages and relatively lower frequency of Severe outages.

**Figure 6.**
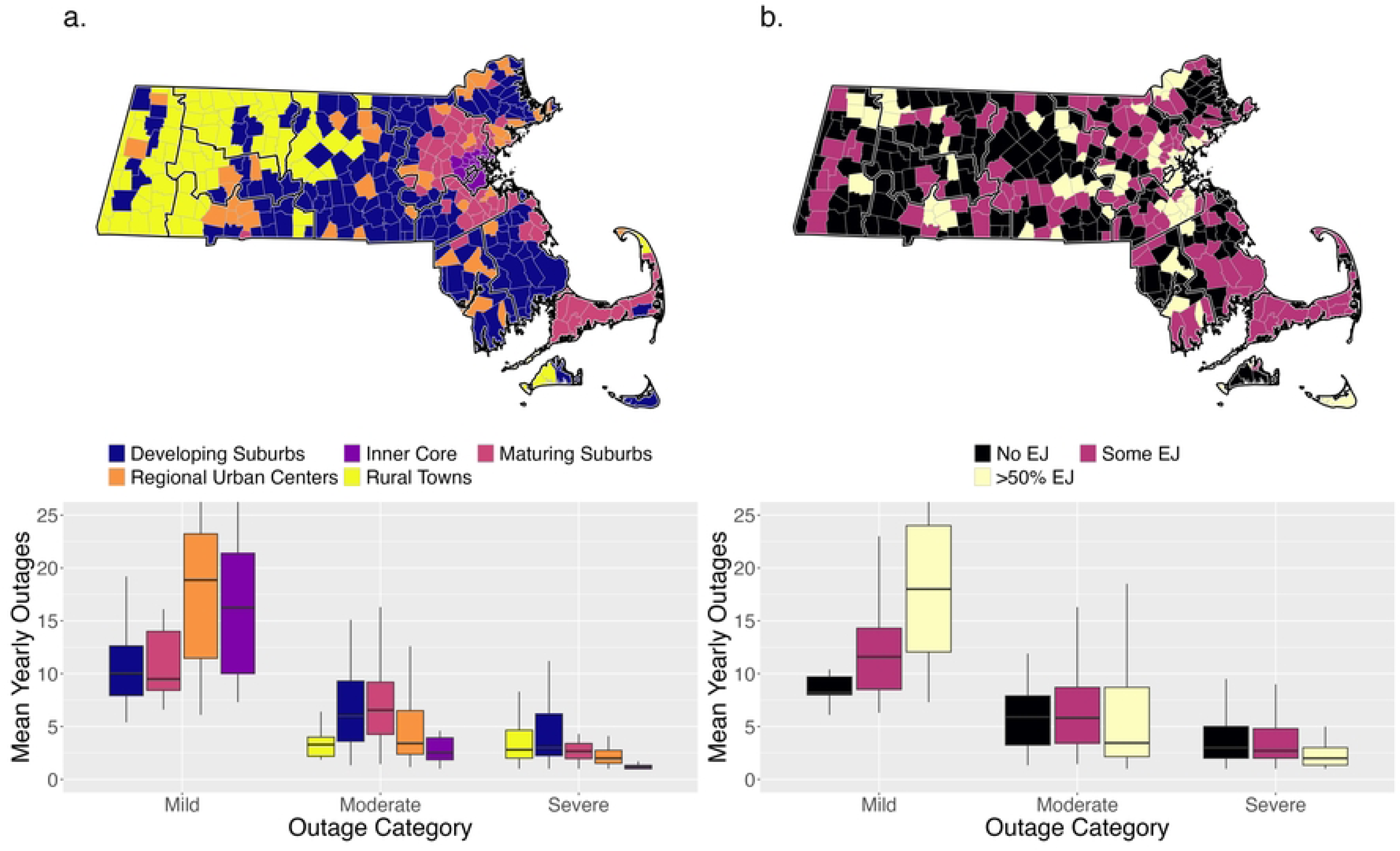
Frequency of power outages by severity and town attributes. Panel (a) shows towns separated by MAPC town classification (the city of Boston is located in the Inner Core); panel (b) shows towns categorized by the fraction of the population that is classified as an Environmental Justice (EJ) population. Towns are outlined in grey, and counties are outlined in black. For ease of comparison, the y-axis has been focused to include the inter-quartile range of each outage severity category within the respective groupings.

## Discussion

Our study leveraged public records from the Massachusetts Department of Public Utilities to develop a daily power outage dataset from 2013 to 2022 at the town-level across the state. We categorized outages by tertile of the number of electrical circuits affected, creating categories for Mild, Moderate, and Severe outages, and describe observed patterns in power outages by community characteristics. We demonstrated seasonal variation in power outages, with the root causes varying both by season and severity. We further demonstrated spatial variation in the average number of power outages, with systematic differences in both magnitude and severity as a function of community type. These data indicate the importance of capturing spatial and temporal variation within studies characterizing power outages, and our methods provide a blueprint for how similar data can be prepared from public records in other states.

### Comparison to other power outage exposure datasets

The dataset created from detailed public records utility data had fine spatial resolution (towns rather than counties) across a long duration (10 years of daily data), with a tiered exposure metric (tertiles of percentage of a town’s circuits on a particular day) and detailed labeling on the suspected cause of each outage. Each of those attributes is valuable for future analyses and has been lacking in many studies to date (**Table 1**).

The importance of spatial resolution can be seen in **Figure 5**. Through town-level analyses, we show that the most urban areas and those with high percentages of Environmental Justice populations have the highest exposure to repeated Mild outages, insight that would not be available through county-level analyses. Such repeated exposure to mild outages has been shown to be correlated with mental health impacts among students at the University of Ghana.^30^ More generally, Mild outages were more associated with equipment failures and planned maintenance rather than tree interference often associated with extreme weather events, indicating differences in both emergency management response and concurrent exposures influencing health. Conversely, more severe outages are seen in more rural and suburban areas, with the associated implications for emergency planning and health risks. Only research in New York State has utilized outage information at finer spatial scales – zip code (spatially approximated by ZIP Code Tabulation Areas, or ZCTAs, **Table 1**); in Massachusetts, there are 539 ZCTAs, meaning that the 351 cities and towns capture a substantial portion of the information that would be available at ZCTA resolution. Furthermore, public health in Massachusetts (and across New England) is largely centered at the municipal rather than county level, making spatially granular analyses all the more important.

The duration of data permits time-series regression of exposures useful for this work and in future study of health impacts related to power outages, potentially in relation to extreme weather events. Using this time-series, we can quantify the increase in outage frequency in March and October, building on similar findings from recent work.^23^ However, we also show that Moderate and Severe outages in Massachusetts are common in July. This may have particular consequences for electricity use that powers air conditioners during summertime. This dataset is among the longest state-level records of daily power outage data available for study (**Table 1**).

Comparisons of our data with prior work illustrate that the choice of metric can have a large impact on the frequency of reported outages. Most studies define a binary variable using a percentile threshold of the percentage of customers in a specific area that are experiencing an outage (**Table 1**). However, these thresholds have vastly different values across states. For example, in several studies in New York,^22–24^ the 50^th^ percentile threshold corresponds to 0.5% of customers in an outage, far lower than in this study in Massachusetts (the 50^th^ percentile in this study represents an outage where 5.8% of circuits are in outage). If we used this threshold in our study, most Mild outages and all Moderate and Severe outages would be counted together in a single category, limiting our capacity to look at heterogeneity in outcomes or develop dose response relationships for health. Later studies in New York have higher thresholds for the 50^th^ and 75^th^ percentile of 2.2% and 1.72%.^14,31^ If any of the New York thresholds were used as cutoffs in this study, the frequency of power outages in Massachusetts would be much higher than reported above. Different from other work, we also include the lowest tertile of exposure – as stated previously, repeated exposure to even low-level outages may have health consequences.

The most common cause of outages across the state was interference from trees, which in many situations may have been downed because of extreme weather events. Foundational work on power outages at the national scale identified extreme weather associated with the majority of outages lasting greater than 8 hours.^1^ Future work can identify the association between this type of outage and extreme weather (e.g., hurricanes), and the types of health outcomes most likely to follow outages associated with each cause type. Continuing to examine the health relevance of varying types of power outages at high spatial and temporal resolution is critical area for future research and advocacy.

### Limitations

Our approach to collecting power outage data had some important limitations to keep in mind when interpreting our results. The ideal exposure metric would be building level – circuit level is a proxy measure that well approximates town population but with the assumption that customers are equally distributed among circuits. Most power outage exposure datasets are aggregated spatially to some extent, and the spatial resolution in this work (city/town scale) is higher than other available datasets for this region. We were unable to extract outage duration, as utility companies did not consistently record this information. Files provided by the utility companies occasionally contained missing or duplicated data for certain towns or months, spelling errors, abbreviations for town names, and irregular spacing. These were remediated to the best of our ability during quality assurance checks. Some circuits were listed in multiple towns, and we assumed that they served equal numbers of customers in each town.

### Future research directions

There are many future directions for research using this novel dataset. A primary application is time series analysis with data pertaining to health outcomes in Massachusetts, particularly looking at weather as a potential effect modifier for health outcomes, as past literature has suggested outages caused by different forms of extreme weather are associated with a higher incidence of health outcomes. Future work should also consider outage causes, as it is possible that certain regions, such as those with aging infrastructure and lower socioeconomic status, could experience more outages caused by equipment failure or malfunction. Future studies on historical procedures’ and policies’ implications for present-day energy injustice should be explored, including on equitable services provided by utility companies and the consequences of electrification on changing power outages.^32^ Additional studies could be conducted on how utility companies have changed over time, including their service areas, mergers with other utility companies, and amount and size of power outages reported. All of these factors can influence town-level outages, and subsequent health impacts. These findings can enable local advocacy and provide ample grounds for future research identifying specific vulnerable populations and evaluating health impacts related to power outages over time. In places where a state-curated dataset is not available, these methods can be used to generate an exposure dataset for power outages useful for emergency planning and epidemiological analyses. The methods here can be replicated to create long time-series of power outage data in other states that have records of such data in distributed formats.

## Conclusion

This study presents a novel, longitudinal, town-level dataset of power outages in Massachusetts from 2013 to 2022, developed using publicly available records from utility providers. The data show clear seasonal patterns and highlight how causes and severity of outages differ by region and community type— critical information for understanding differential vulnerabilities and guiding equitable emergency preparedness and infrastructure planning.

Our methodology offers a replicable approach to constructing granular power outage datasets from administrative records, which can support epidemiologic research and climate adaptation planning in other regions. The dataset’s resolution and duration enable investigations into the health impacts of outages, particularly in the context of climate-related extreme weather events. Future research should build on this foundation to examine the interaction of outage types, causes, and health outcomes across vulnerable populations. By making this data infrastructure more accessible and actionable, we move toward more just and informed responses to the growing public health threat of power outages.

## Data availability

Sample code is available on Github: https://github.com/cmilando/DPU_poweroutage_demo. The complete analytical data have been entered into a figshare associated with this manuscript.

## Conflicts of interest

We have no conflicts of interest to declare.

## Acknowledgements

This project was partially funded by the United States Environmental Protection Agency (EPA) under assistance agreement RD-84048001 to Boston University, until the assistance agreement was terminated in May 2025 for no longer effectuating agency priorities. We thank Flor Amaya in the City of Chelsea, MA Department of Public Health, for suggesting we examine power outages. The contents of this document do not necessarily reflect the views and policies of the EPA, nor do they endorse trade names or recommend the use of commercial products mentioned in this document.

## Supplemental captions

**Table S1.** Words ascribed to each cause type category

**Table S2.** Annual outages by town. NA indicates no outages that year.

**Table S3.** Regression coefficients

## Notes

### Competing Interest Statement

The authors have declared no competing interest.

### Funding Statement

Yes

